# Neurodegenerative Plasma Biomarkers for Prediction of Hippocampal Atrophy in Older Adults with Suspected Alzheimer’s Disease in Kinshasa, Democratic Republic of Congo

**DOI:** 10.1101/2024.09.03.24313019

**Authors:** Jean Ikanga, Kharine Jean, Priscilla Medina, Saranya Sundaram Patel, Megan Schwinne, Emmanuel Epenge, Guy Gikelekele, Nathan Tshengele, Immaculee Kavugho, Samuel Mampunza, Lelo Mananga, Charlotte E. Teunissen, Anthony Stringer, Julio C. Rojas, Brandon Chan, Argentina Lario Lago, Joel H. Kramer, Adam L. Boxer, Andreas Jeromin, Bernard Hanseeuw, Alden L. Gross, Alvaro Alonso

**Author notes:** Corresponding author. Please send general questions about the study to Dr. Jean Ikanga. Jean N. Ikanga, Ph.D., Department of Rehabilitation Medicine, Emory University, 1441 Clifton Rd NE, Atlanta, GA 30322, USA.

## Abstract

**Objective:** The hippocampus is one of the first brain structures affected by Alzheimer’s disease (AD), and its atrophy is a strong indicator of the disease. This study investigates the ability of plasma biomarkers of AD and AD-related dementias— amyloid-β (Aβ42/40), phosphorylated tau-181 (p-tau181), neurofilament light (NfL), and glial fibrillary acidic protein (GFAP)—to predict hippocampal atrophy in adult individuals in Kinshasa, Democratic Republic of Congo (DRC).

**Methods:** Eighty-five adult individuals (40 healthy and 45 suspected AD) over 65 years old were evaluated using the Community Screening Instrument for Dementia and Alzheimer’s Questionnaire (AQ). Core AD biomarkers (Aβ42/40 and p-tau181) and non-specific neurodegeneration biomarkers (NfL, GFAP) were measured in blood samples collected at the study visit. Hippocampal volumes were measured using magnetic resonance imaging (MRI). General linear regression was used to evaluate differences in biomarker concentrations by neurological status. Logistic regression models were used to create receiver operating characteristic curves and calculate areas under the curve (AUCs) with and without clinical covariates to determine the ability of biomarker concentrations to predict hippocampal atrophy. Plasma biomarkers were used either individually or in combination in the models.

**Results:** Elevated p-tau181 was associated with left hippocampal (LH) atrophy p= 0.020). Only higher p-tau181 concentrations were significantly associated with 4.2-fold increased odds [OR=4.2 (1.5-18.4)] of hippocampal atrophy per standard deviation. The AUC of plasma biomarkers without clinical covariates to discriminate LH, RH, and total hippocampal (TH) or both hippocampi atrophy ranged between 90% to 94%, 76% to 82%, and 85% to 87%, respectively. The AUC of models including clinical covariates and AD biomarkers used in combination to discriminate LH, RH, and TH ranged between 94%-96%, 81%-84%, and 88%-90%, respectively.

**Conclusion:** These results indicate that, consistent with studies in other settings, core AD plasma biomarkers can predict hippocampal atrophy in a population in Sub-Saharan Africa.

## Introduction

Alzheimer’s disease (AD) is a progressive neurodegenerative disorder that is associated with hippocampal atrophy.^1^ AD is notable for cognitive decline predominantly impacting memory functioning alongside deficits in other neurocognitive domains.^1^ Ongoing research of AD pathology has expanded the number of fluids (e.g., cerebral spinal fluid [CSF], blood) biomarkers utilized in the screening, diagnosis, and monitoring of AD,^2^ including the core biomarkers of AD [ratio of amyloid-β (Aβ42/40), phosphorylated tau-181 (p-tau181)], and non-specific biomarkers to AD [neurofilament light (NfL), and glial fibrillary acidic protein (GFAP)]. These studies have shown a positive and stronger correlation between CSF and plasma mostly with Aβ42 and Aβ40 levels.^2–7^

Amyloid and tau deposition, axonal damage and astrocyte activation are integral to neurodegenerative cerebral atrophy.^8^ Aβ accumulation pathway begins with the cortices, allocortical regions, midbrain, cerebellum, and brain stem.^9^ In contrast, tau accumulation starts in mesial temporal lobes and continues to the allocortical and neocortical regions of the temporal lobe, subsequently extending to the parietal lobe, occipital, prefrontal areas, premotor areas, and finally into the neocortical primary fields.^10^

The hippocampus is one of the first brain structures affected with atrophy in AD.^1,11^ Hippocampus is known for its key roles in memory consolidating, which is a key deficit in AD patients.^12^ Therefore, hippocampal atrophy is considered as an important clinical feature in AD. The atrophy of hippocampus is a strong indicator of AD, and the rate of its shrinkage is used to predict the progression of AD.^13^ Currently, the literature is mixed regarding the association between plasma AD biomarkers and hippocampal volume. In participants from the Baltimore Longitudinal Study of Aging (BLSA), baseline plasma Aβ42/40 ratio, GFAP, NfL, and p-tau181 were not associated with baseline hippocampal volume.^2^ However, for those with lower amyloid burden, greater baseline ptau-181 was associated with accelerated decline in hippocampal volume, possibly reflecting greater impact during the earlier course of AD pathology. In another study examining four older adult cohorts across the United States, plasma GFAP was not associated with hippocampal volume.^3^ In contrast, one study found that plasma AD biomarkers (p-tau 181, Aβ42/40, NfL) predicted hippocampal atrophy in a sample of cognitively-healthy older adults.^4^ Additionally, those predictions were specific to AD, as there were no associations found in a sample of non-AD individuals.^4^

Overall, most AD biomarker studies have primarily been conducted with Western populations samples made up of predominantly White individuals of European ancestry. However, non-White populations may differ from White populations in terms of cognitive reserve, cultural background, lifestyle, and genetics. Specifically, cognitive testing is not well standardized for sub-Saharan populations and costly AD biomarkers, such as CSF or PET, are little available. Thus, understanding the role of plasma AD biomarkers within non-White populations is crucial for generalizability of AD clinical research, especially in relation to emerging diagnostic and therapeutic tools. There are few studies on the association of core AD biomarkers (Aβ and p-tau) and hippocampal atrophy in culturally diverse populations. For example, in an enthnoracially diverse sample of community dwelling older adults, hippocampal volume was found to be a significant mediator between Aβ42/40 and NfL on baseline episodic memory and executive function measures.^14^ In a Singaporean cohort with varying cognitive status, p-tau181, p-tau181/t-tau, Aβ42/40, and p-tau 181/Aβ42 ratios were all significantly associated with hippocampal volume.^15^ Similar associations were seen in a primarily Hispanic sample (>60% Hispanic participants), where hippocampal volume was significantly related to plasma NfL in individuals with AD, but not for those who were considered cognitively normal.^16^

The current study aims to investigate ability of AD core plasma biomarkers (Aβ 42/40, p-tau181) and non-specific AD biomarkers (NfL and GFAP) to discriminate the severity of hippocampal atrophy in adult individuals in Kinshasa, Democratic Republic of the Congo (DRC), in Sub-Saharan Africa (SSA). Although comparing left versus right hippocampal atrophy is not the central focus of this study, we will consider a lateralization approach. Our main goal is to describe the clinical value of these biomarkers in this novel population. This opens the door to more in-depth explorations of their clinical value and the examination of the left-sided vulnerability theory in dementia patients within this African population. We hypothesized that core (Aβ42/40 and p-tau181) and non-specific (NfL and GFAP) plasma biomarkers of AD will be associated with hippocampal atrophy in this sample. Given the pathophysiological impact and the usefulness of p-tau181 in the diagnosis of AD pathology,^17^ we hypothesized that p-tau181 will have greater discriminatory ability of hippocampal atrophy than other plasma biomarkers. Finally, we predicted that Aβ42/40, NfL, and GFAP will demonstrate adequate to good sensitivity to discriminate between individuals with and without hippocampal atrophy.

## METHODS

### Study population

Participants of this study are community-dwellers from Kinshasa, DRC, selected from our previous study.^18^ Participants were included if they were at least 65 years or older, had a family member or close friend to serve as an informant, and were fluent in French or Lingala. We excluded participants who had history of schizophrenia, neurological disease other than dementia, or other medical conditions potentially affecting the central nervous system (CNS). To establish neurological status in the absence of established diagnostic criteria for AD in SSA, we screened participants using the Alzheimer’s Questionnaire (AQ)^19^ and the Community Screening Instrument for Dementia (CSID).^20^ The AQ was used to assess activities of daily living and symptoms of AD in participants.^19^ The CSID Questionnaire, which is extensively used in many SSA dementia studies,^20^ was used to screen cognitive abilities. Based on cognitive and functional deficits per the Diagnostic and Statistical Manual of Mental Disorders, Fifth Edition, Text Revision (DSM-5-TR) diagnostic criteria,^21^ we used Congo-Brazzaville cut-offs of CSID, the closest city from Kinshasa, to classify participants.^22^ Similar to our prior study,^23^ participants were classified using CSID and AQ scores (see Figure 1), which yielded 4 groups: dementia, mild neurocognitive disorder (MND), subjective cognitive impairment (SCI), and healthy control (HC), i.e., normal cognition. Due to our case-control design, we excluded participants with MCI and SCI. A panel consisting of a neurologist, psychiatrist and neuropsychologist reviewed screening tests, clinical interview, and neurological examination of subjects, of whom 56 were confirmed with a diagnosis of dementia and 58 were considered HC. Of these 114 participants, 29 refused to provide blood samples, leaving 85 participants (75%) in whom plasma biomarkers were obtained (44 dementia and 41 HC). Written informed consent was obtained prior to participants’ undergoing any study procedures. Participants were financially compensated for their time. The procedures were approved by the Ethics Committee/Institutional Review Boards of the University of Kinshasa and Emory University.

**Figure 1.**
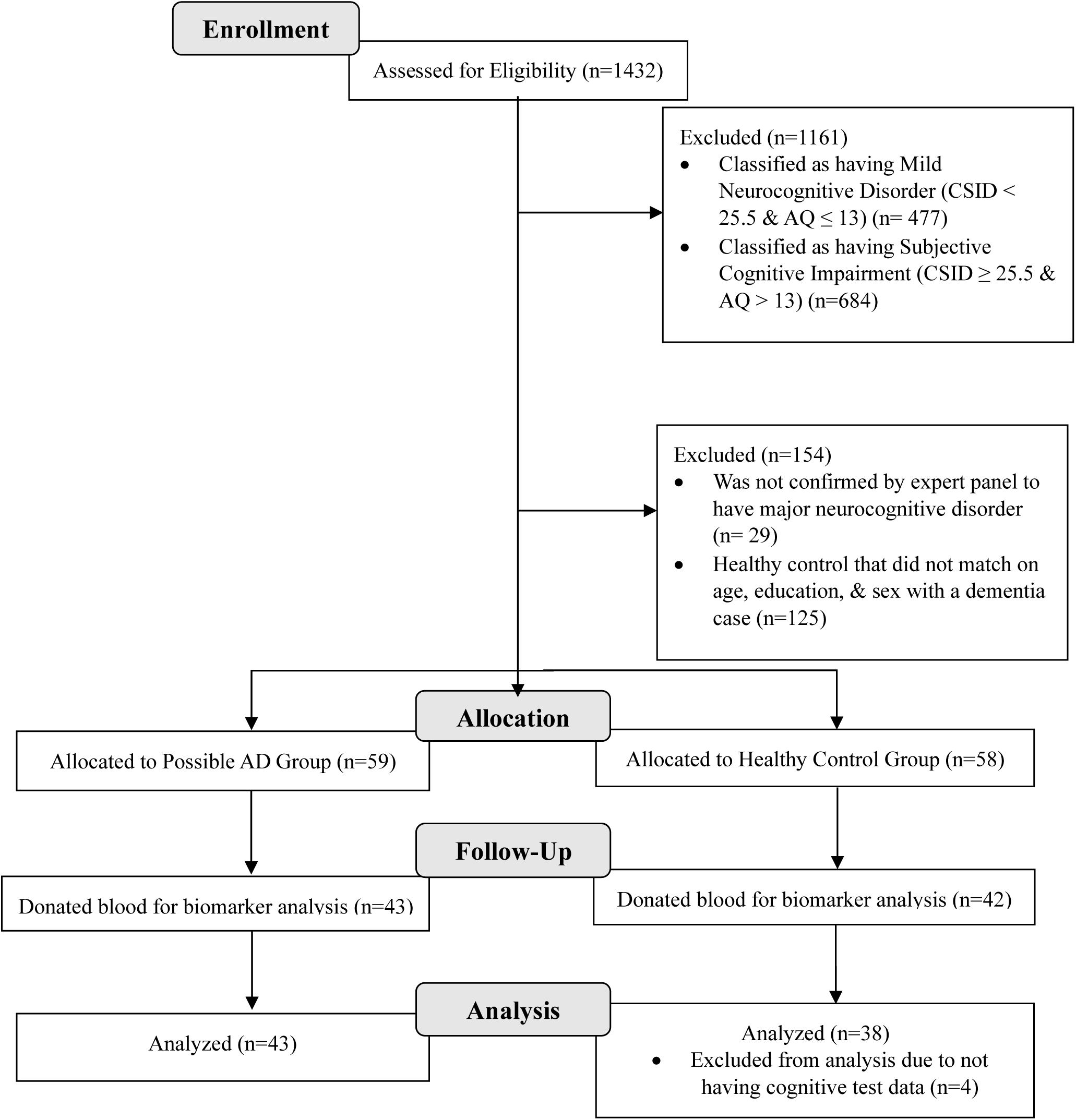
Flow Chart of Recruitment Status from those assessed for eligibility at enrollment (n=1432) to the individuals that were allocated to the dementia or control group and analyzed (n=85)

### Procedure

Participants underwent a comprehensive clinical evaluation, which included cognitive testing, self-report questionnaires, and standard psychiatric and neurological assessments. An expert panel consisting of a neurologist, psychiatrist, and neuropsychologist diagnosed participants with dementia or classified them as healthy controls (HC). Subjects were interviewed to gather demographic, socioeconomic, and medical history information. Subsequently, blood samples were collected by a phlebotomist at the Medical Center of Kinshasa (CMK). The sample collection protocol and quantification of fluid biomarkers are detailed below.

### Measures

#### Plasma biomarkers

Blood samples were drawn in the CMK blood laboratory by venipuncture into dipotassium ethylene diamine tetra acetic acid (K_2_ EDTA) tubes. Samples were centrifuged within 15 minutes at 1800 g house temperature, and 5 mL of plasma was aliquoted into 0.5 mL polypropylene tubes and stored initially at −20° C for less than a week and then moved to a −80 °C freezer for longer term storage at a CMK laboratory.^24^ These aliquots were shipped frozen on dry ice to Emory University.

Plasma biomarker concentrations were measured using commercially available Neurology 4-PLEX E (Aβ40, Aβ42), P-tau181 (P-tau181 v2; l) Quanterix kits for the Simoa HD-X platform (Billerica, MA) at the University of San Francisco. The instrument operator was blinded to clinical variables. All analytes were measured in duplicate. For Aβ40 and Aβ42, all samples were measured above the lower limit of quantification (LLOQ) of 1.02 pg/mL and 0.378 pg/mL. The average coefficient of variation (CV) for Aβ40 and Aβ42 were 6.0% and 6.5%. For P-tau181, all samples were measured above the kit lower limit of quantification (LLOQ) of 0.085 pg/mL, with an average CV of 11.6%.

### Neuroimaging

All subjects were imaged on a 1.5 Tesla MRI unit (Siemens, Magneton Sonata) scanner at HJ Hospitals in Kinshasa using the same standardized imaging acquisition protocol based on the Alzheimer’s Disease Research Center protocol of Emory University. This consisted of sagittal volumetric T1-weighted (MPRAGE), coronal T2-weighted, and axial diffusion-weighted, T2-weighted, and T2-FLAIR sequences. Typical acquisition parameters for the MPRAGE sequence were TR = 2200 ms, minimum full TE, TI = 1000 ms, flip angle = 8°, FOV = 25 cm, with a 192 × 184 acquisition matrix, yielding a voxel size of approximately 1.25 × 1.25 × 1.2 mm.^25^ Images were reviewed by an experienced neuroradiologist. White matter hyperintensity was graded according to the age-related white matter changes (ARWMC) scale.^26^ Number of chronic brain parenchymal microhemorrhages were recorded. Lobar volume loss pattern of the brain was assessed. MPRAGE images were reoriented into the oblique coronal plane orthogonal to the principal axis of the hippocampal formation, and medial temporal lobe atrophy (MTLA) and entorhinal cortex atrophy (EriCa) scores were assessed.^27^ Finally, the presence or absence of any additional abnormalities was noted, and patients were excluded if neuroimaging evidence indicated an etiology other than probable AD (e.g., presence of a brain tumor).

#### Quantitative volumetric analysis using Freesurfer

The 3D T1w images were segmented using Freesurfer (v.6, MGH, MA), which includes a full processing stream for MR imaging data that involves skull-stripping, bias field correction, registration, and anatomical segmentation as well as cortical surface reconstruction, registration, and parcellation. Regional brain volume for both cortical and subcortical brain regions were calculated. The left and right hippocampal (LH, RH) volume was averaged. Interindividual variation in head size were accounted for in further statistical analysis by controlling for total intracranial volume.

### Statistical Analyses

Statistical analyses were performed using SAS and R Version 4 statistical softwares. Descriptive statistics for continuous, normally distributed variables are presented as mean ± standard deviation (SD), while categorical variables are expressed using counts and proportions. Winsorization of plasma biomarkers to the 95th percentile was applied to limit the effect of extreme outliers. Biomarkers were standardized by subtracting the mean biomarker value of the sample and dividing by the sample’s plasma biomarker standard deviation. Standardized hippocampal volumes and biomarkers were obtained for the left hippocampus (LH), right hippocampus (RH), and total hippocampal (TH) volume. We calculated the differences in hippocampal volumes based on cognitive status and defined hippocampal atrophy using established cutoffs:^28^

- ≥ 3000 mm^3^ or < 3000 mm^3^ to define normal or atrophy for LH and RH, respectively.
- ≥ 6000 mm^3^ or < 6000 mm^3^ to define normal or atrophy for TH, respectively.

Based on these cutoffs, we calculated the prevalence of LH, RH and TH atrophy.

General linear regression, adjusting for age, sex, and education, was used to evaluate differences in biomarkers by neurological status. Logistic regression was conducted with left, right, or total hippocampal volume <3000 or <6000 mm^3^ as outcome variable (atrophy) and biomarker as primary independent variable, controlling for age, gender, education, geriatric depression scale (GDS) score, and intracranial volume.

Biomarkers are standardized. Additionally, logistic regression was conducted to create receiver operating characteristic (ROC) curves and calculate areas under the curve (AUCs) to evaluate the ability of plasma biomarkers (Aβ42/40, p-tau 181, NfL, GFAP) to predict hippocampal atrophy, controlling for age, education, gender, depression score, and intracranial volume. Cutoff scores for each plasma biomarker were determined based on optimal sensitivity and specificity for detecting hippocampal atrophy, using the value that maximized Youden’s index (sensitivity + specificity – 100). We used the ROC-AUC categories defined by Hosmer and colleagues (Hosmer et al., 2013), which classify values as follows: <0.600 as “failure”, 0.600 to 0.699 as “poor”, 0.700 to 0.799 as “fair”, 0.800 to 0.899 as “good”, and ≥0.900 as “excellent.”

## RESULTS

Demographic data, cognitive screening scores, hippocampal volumes, and plasma biomarker characteristics stratified by neurological status are presented in Table 1. There were no significant differences in sex between groups. Education level was lower in participants with suspected dementia. There were significant differences in cognitive screening scores used in distinguishing neurological status, with HC having higher scores than those with suspected AD. There was trend of differences in LH, RH, and TH volumes between HC and suspected AD, with suspected AD showing lower hippocampal volumes. NfL and GFAP also differed significantly by neurological status after controlling for age, gender, and education, with suspected AD having higher concentration levels than HC (Table 1). The difference between the TH volume of HC and suspected AD was 367 mm^3^. The difference between the LH and RH volumes of HC was 30 mm^3^, while the difference between LH and RH volumes of suspected AD was 29 mm^3^. Frequency of LH, RH and TH atrophy was 32.5 %, 34.2%, and 33.3%, respectively.

**Table 1.** Demographic and Clinical Characteristics of Individuals with Neurological Status.

| | Healthy,<br>$\mu$ ($\sigma$),<br>(n=29) | Suspected AD,<br>$\mu$ ($\sigma$),<br>(n=47) | $\beta_1^*$<br>(95% CI) | p-value |
| --- | --- | --- | --- | --- |
| Age (years) | 71.5 (8.7) | 73.0 (7.7) | -0.28<br>(-3.8, 3.2) | 0.87 |
| Male (n,%)† | 18 (64%) | 22 (47%) | -0.11<br>(-1.4, 1.1) | 0.86 |
| Education (years) | 11.0 (5.0) | 8.2 (5.3) | -1.4<br>(-3.2, 0.4) | 0.12 |
| GDS Score | 4.0 (2.4) | 8.1 (3.8) | 4.1<br>(2.4, 5.7) | <.0001 |
| CSID | 32.5 (2.6) | 20.0 (5.3) | -11.8<br>(-14.1, -9.6) | <.0001 |
| AQ | 2.5 (2.1) | 18.5 (3.7) | 15.9<br>(14.3, 17.5) | <.0001 |
| Brain Structures |  |  |  |  |
| Total Hippocampus (mm<sup>3</sup>) | 6824 (858) | 6062 (1036) | -0.47<br>(-0.09, 0.15) | 0.058 |
| Left Hippocampus (mm<sup>3</sup>) | 3426 (453) | 3046 (560) | -0.023<br>(-0.05, 0.22) | 0.073 |
| Right Hippocampus (mm<sup>3</sup>) | 3398 (440) | 3017 (560) | -0.023<br>(-0.05, 0.39) | 0.092 |
| ICV (mm<sup>3</sup>) | 1439986<br>(159015) | 1415062<br>(261524) | -41377<br>(-154337, 71584) | 0.47 |
| Biomarkers |  |  |  |  |
| A$\beta_{42}$ (pg/ml) | 3.2 (1.8) | 3.6 (2.0) | 0.24<br>(-0.9, 1.4) | 0.68 |
| A$\beta_{40}$ (pg/ml) | 51.5 (37.2) | 71.9 (49.1) | 15.1<br>(-11.5, 41.7) | 0.26 |
| A$\beta_{42/40}$ | 0.08 (0.04) | 0.06 (0.03) | -0.01<br>(-0.04, 0.007) | 0.18 |
| p-tau 181 (pg/ml) | 1.9 (1.3) | 2.7 (2.2) | 0.51<br>(-0.60, 1.6) | 0.36 |
| NfL (pg/ml) | 26.4 (11.8) | 66.5 (43.3) | 39.5<br>(20.0, 59.1) | 0.0002 |
| GFAP (pg/ml) | 136 (48) | 238 (139) | 98.5<br>(38.2, 158.7) | 0.0018 |

Biomarkers are not standardized. Data is for individuals in this population that have data on hippocampal volume. ICV = Intracranial Volume, CSID = Community Screening Instrument for Dementia, AQ = Alzheimer's Questionnaire, A $\beta_{42/40}$ = ratio of Amyloid- $\beta$ 42 and 40 peptides, p-tau = Phosphorylated tau, NfL = Neurofilament Light, GFAP = Glial Fibrillary Acidic Protein.
\* Represents the linear effect of the measure as the outcome against neurological status, controlling for age, gender, and education. Brain structures also control for GDS and Intracranial Volume.
† Represented as n (%) and logistic regression is utilized

Table 2 presents the association between hippocampal atrophy and standardized plasma biomarkers. The volume of the left hippocampus (LH) was significantly associated with p-tau181 concentration. Higher p-tau181 concentrations were linked to a 4.2-fold increase in the odds of hippocampal atrophy per standard deviation. However, the volumes of both the right hippocampus (RH) and the total hippocampus (TH) did not show any significant associations with other standardized biomarkers see Table 2).

**Table 2.** Association between Left, Right, and Total Hippocampus Atrophy and Standardized Biomarkers.

| | **LH-,**<br>$\mu$ ($\sigma$),<br>(n=51) | **LH+,**<br>$\mu$ ($\sigma$),<br>(n=25) | **Odds Ratio\***<br>(95% CI) | **p-value** |
| --- | --- | --- | --- | --- |
| A$\beta_{42/40}$ | 0.17 (1.05) | -0.33 (0.82) | 0.45 (0.14, 1.1) | 0.12 |
| p-tau181 (pg/ml) | -0.35 (0.62) | 0.65 (1.24) | 4.2 (1.5, 18.4) | 0.020 |
| NfL (pg/ml) | -0.14 (0.95) | 0.28 (1.1) | 1.9 (0.65, 7.0) | 0.26 |
| GFAP (pg/ml) | -0.34 (0.71) | 0.65 (1.16) | 1.2 (0.43, 3.0) | 0.74 |
| *Right Hippocampus and Standardized Biomarkers* |  |  |  |  |
| | **RH-,**<br>$\mu$ ($\sigma$),<br>(n=49) | **RH+,**<br>$\mu$ ($\sigma$),<br>(n=27) | **Odds Ratio\***<br>(95% CI) | **p-value** |
| A$\beta_{42/40}$ | 0.18 (1.05) | -0.37 (0.79) | 0.55 (0.22, 1.1) | 0.13 |
| p-tau181 (pg/ml) | -0.27 (0.77) | 0.54 (1.20) | 1.9 (0.97, 4.0) | 0.077 |
| NfL (pg/ml) | -0.10 (1.01) | 0.20 (0.98) | 1.3 (0.59, 3.0) | 0.51 |
| GFAP (pg/ml) | -0.21 (0.78) | 0.41 (1.25) | 1.3 (0.64, 2.8) | 0.46 |
| *Total Hippocampus and Standardized Biomarkers* |  |  |  |  |
|  | **TH-,**<br>**μ (σ),**<br>**(n=49)** | **TH+,**<br>**μ (σ),**<br>**(n=27)** | **Odds Ratio\***<br>**(95% CI)** | **p-value** |
| Aβ<sub>42/40</sub> | 0.20 (1.1) | -0.35 (0.80) | 0.47 (0.18, 1.0) | 0.078 |
| p-tau 181 (pg/ml) | -0.30 (0.62) | 0.52 (1.29) | 2.0 (0.93, 4.8) | 0.10 |
| NfL (pg/ml) | -0.12 (0.97) | 0.20 (1.04) | 1.6 (0.62, 4.6) | 0.34 |
| GFAP (pg/ml) | -0.32 (0.73) | 0.53 (1.17) | 1.5 (0.65, 3.5) | 0.36 |

\* Logistic regression model with left, right, or total hippocampal volume <3000 (right, left) or <6000 (total) mm<sup>3</sup> as outcome variable (atrophy) and biomarker as primary independent variable, controlling for age, gender, education, GDS score, and intracranial volume. Biomarkers are standardized.

Table 3 presents the cutoffs, sensitivity, specificity, and accuracy of plasma biomarkers in discriminating LH atrophy. Sensitivity varied from poor (50%) to fair (79%), with Aβ42/40 and p-tau181 showing the highest sensitivity. Specificity was good, ranging from 56% to 85%. The crude AUC of plasma biomarkers for predicting LH atrophy ranged from 53% to 77%. The AUC of the adjusted model, which included plasma biomarkers and covariates, was excellent, ranging from 90% to 94%, with p-tau181 having the highest AUC and GFAP the lowest. Figure 2 illustrates the various AUCs of plasma biomarkers in discriminating LH atrophy. We also developed models using a combination of plasma biomarkers to assess their ability to discriminate LH atrophy. The AUC of the adjusted model, which included covariates and combined plasma biomarkers, was excellent, ranging from 94% to 96%. The AUC of a model including only covariates (without any biomarkers) to discriminate LH atrophy was 85% (see Table 3).

**Figure 2.**
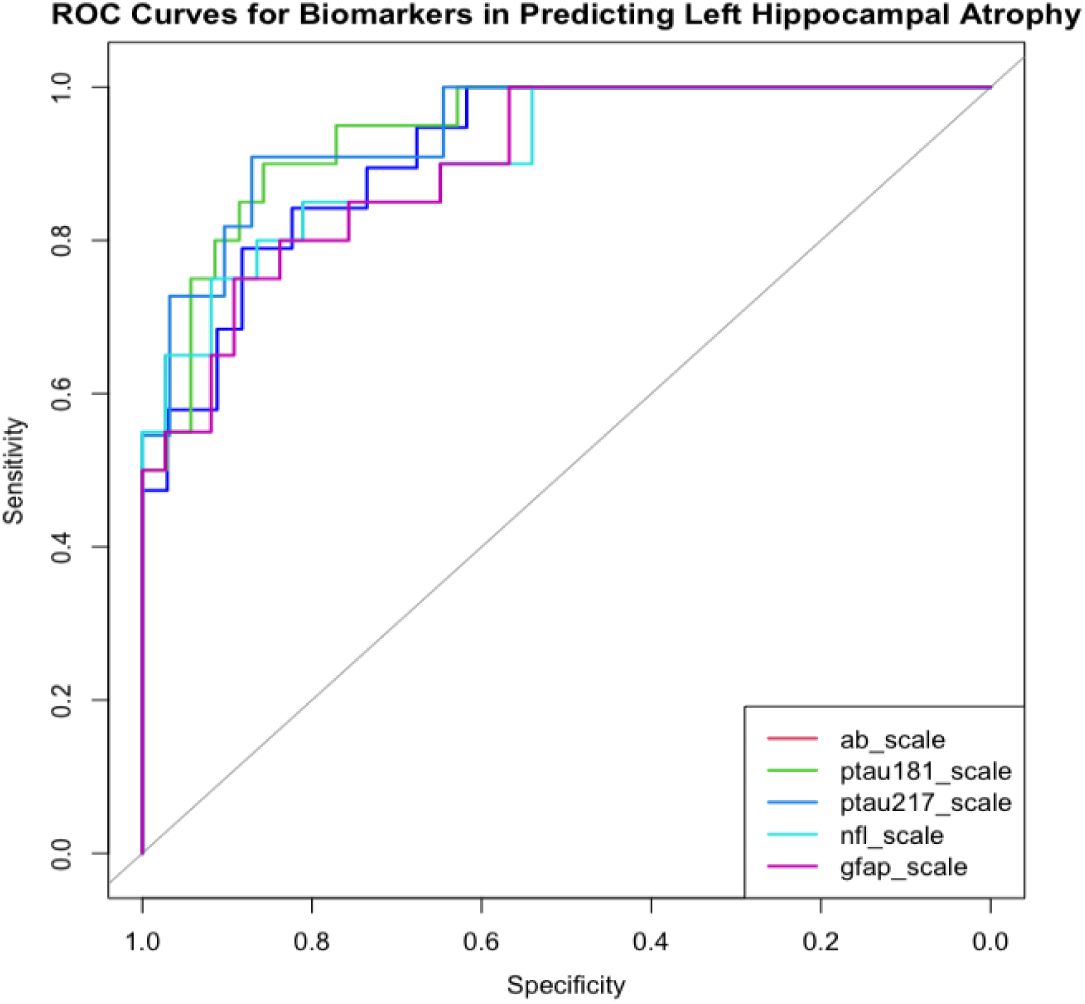
AUC discriminations of plasma biomarkers predicting left hippocampal atrophy

**Table 3.**
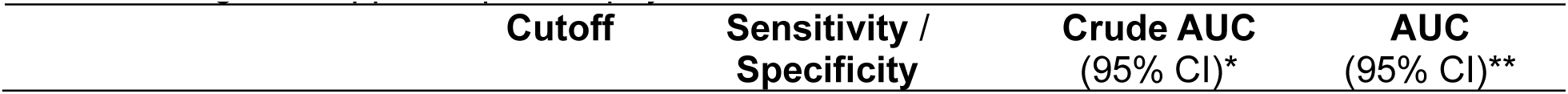
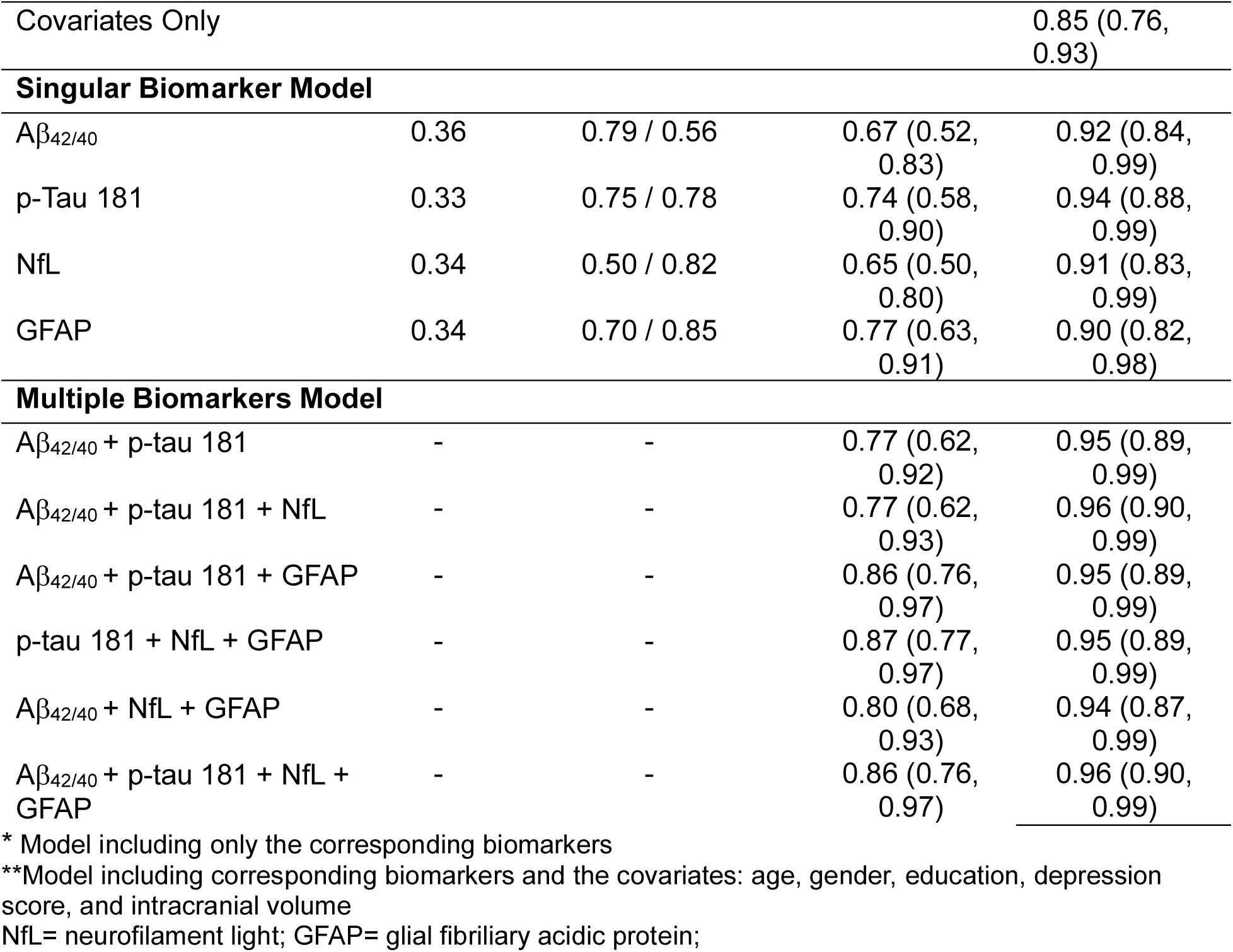
Sensitivity, Specificity, and Accuracy of Standardized Plasma Biomarkers in Discriminating Left Hippocampal Atrophy.

Table 4 presents the cutoffs, sensitivity, specificity, and discrimination ability of plasma biomarkers for RH atrophy. The sensitivity of plasma biomarkers to discriminate RH atrophy varies between 45% and 79%, with GFAP showing the lowest and p-tau 181 the highest sensitivity. Specificity ranges between 63% and 87%. The crude AUC of plasma biomarkers for discriminating right hippocampal atrophy varied between 64% and 70%. The AUC of the adjusted model, which includes biomarkers and covariates, ranged from 76% to 82%, with p-tau 181 showing the highest predictive ability. Figure 3 illustrates the various AUCs of plasma biomarkers in discriminating RH atrophy. The AUC of a covariates-only model for RH atrophy prediction was 78.0%. The AUC of the combination of biomarkers to discriminate RH atrophy varied between 81% and 86%. The presence of core biomarkers (Aβ42/40 and p-tau 181) had a greater impact on the model (see Table 4).

**Figure 3.**
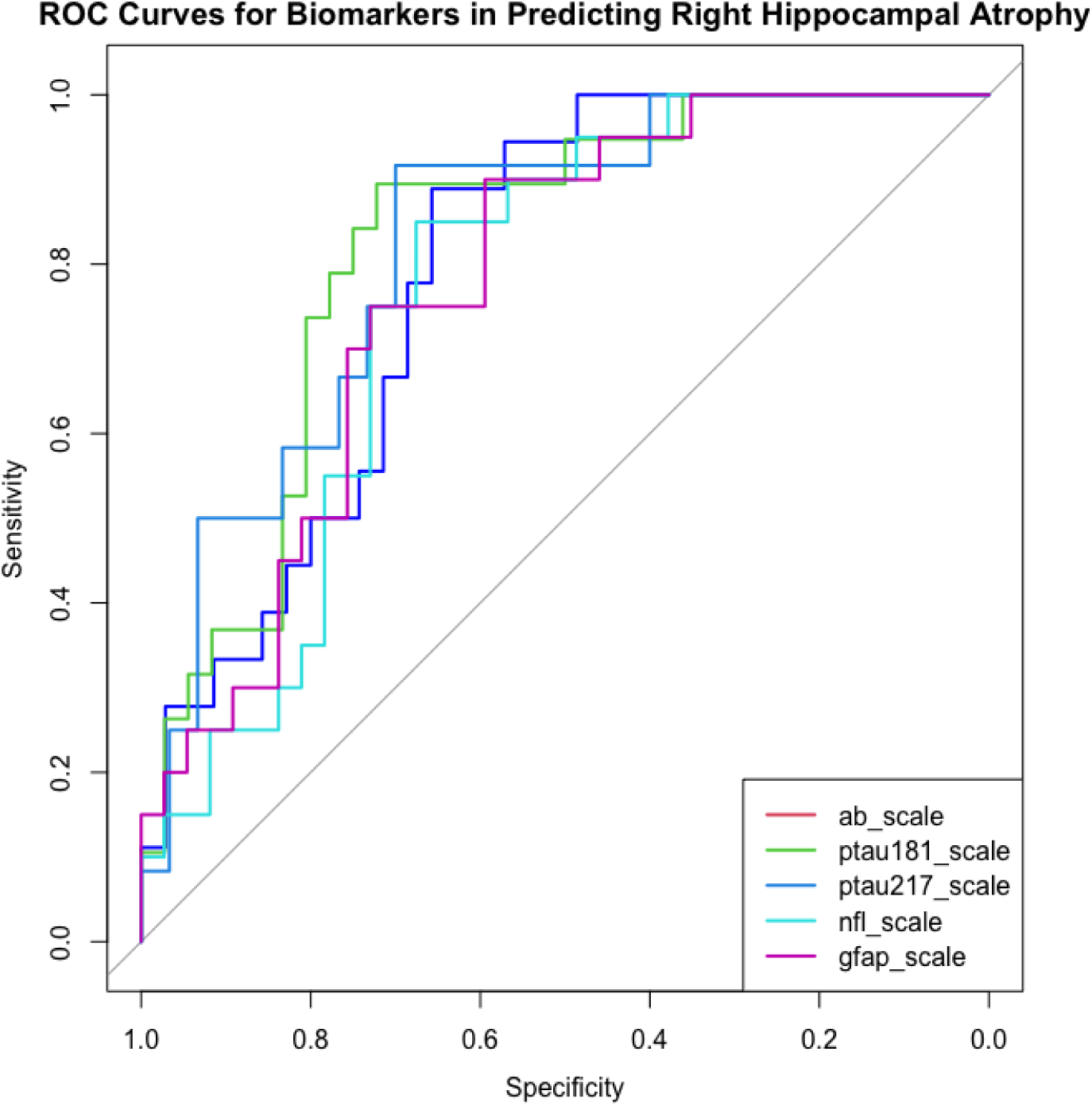
AUC discriminations of plasma biomarkers predicting right hippocampal atrophy

**Table 4.** Sensitivity, Specificity, and Accuracy of Plasma Biomarkers in Discriminating Right Hippocampal Atrophy.

| **Biomarker** | **Cutoff** | **Sensitivity / Specificity** | **Crude AUC (95% CI)\*** | **AUC (95% CI)\*\*** |
| --- | --- | --- | --- | --- |
| Covariates Only |  |  |  | 0.78 (0.68, 0.89) |
| **Singular Biomarker Model** |  |  |  |  |
| A$\beta_{42/40}$ | 0.36 | 0.72 / 0.73 | 0.70 (0.55, 0.85) | 0.79 (0.67, 0.91) |
| p-Tau 181 | 0.27 | 0.79 / 0.63 | 0.70 (0.55, 0.86) | 0.82 (0.71, 0.93) |
| NfL | 0.32 | 0.65 / 0.64 | 0.64 (0.49, 0.79) | 0.76 (0.64, 0.89) |
| GFAP | 0.37 | 0.45 / 0.87 | 0.64 (0.48, 0.80) | 0.77 (0.65, 0.89) |
| **Multiple Biomarkers Model** |  |  |  |  |
| A$\beta_{42/40}$ + p-tau 181 | - | - | 0.74 (0.59, 0.89) | 0.84 (0.73, 0.95) |
| A$\beta_{42/40}$ + p-tau 181 + NfL | - | - | 0.75 (0.60, 0.91) | 0.83 (0.72, 0.94) |
| A$\beta_{42/40}$ + p-tau 181 + GFAP | - | - | 0.74 (0.59, 0.90) | 0.83 (0.72, 0.94) |
| p-tau 181 + NfL + GFAP | - | - | 0.75 (0.59, 0.90) | 0.82 (0.71, 0.93) |
| A$\beta_{42/40}$ + NfL + GFAP | - | - | 0.72 (0.57, 0.87) | 0.81 (0.70, 0.93) |
| A$\beta_{42/40}$ + p-tau 181 + NfL + GFAP | - | - | 0.74 (0.59, 0.90) | 0.83 (0.72, 0.94) |

\* Model including only the corresponding biomarkers
\*\*Model including corresponding biomarkers and the covariates: age, gender, education, depression score, and intracranial volume
NfL= neurofilament light; GFAP= glial fibrillary acidic protein;

Table 5 presents the cutoffs, sensitivity, specificity, and discriminative ability of plasma biomarkers to predict TH atrophy. The sensitivity of plasma biomarkers to discriminate TH atrophy ranged from 50% to 80%, with Aβ_42/40_ showing the highest sensitivity and GFAP the lowest. Specificity varied between 57% and 92%. The crude AUC of plasma biomarkers for discriminating TH atrophy ranged from 63% to 73%. The AUC of the adjusted model, which includes biomarkers and covariates, ranged from 85% to 87%, with p-tau 181 having the highest AUC. Figure 4 illustrates the various AUCs of plasma biomarkers in discriminating TH atrophy. In this model, the AUC of the covariates-only model was 85%. Similar to LH and RH, the AUC in both crude and adjusted models increased with the number of biomarkers included. The AUC varied between 88% and 90% (see Table 5).

**Figure 4.**
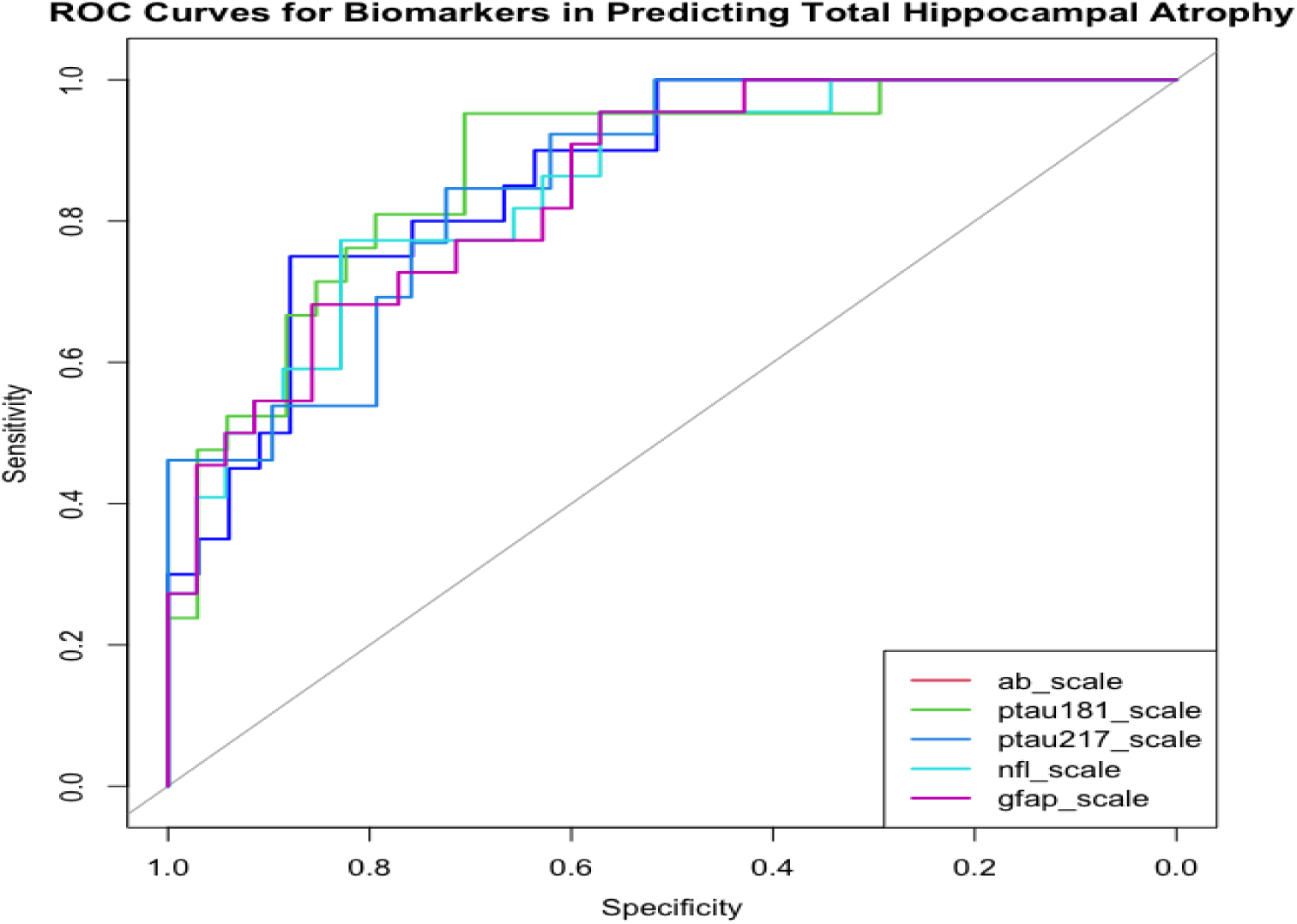
AUC discriminations of plasma biomarkers predicting total hippocampal atrophy

**Table 5.** Sensitivity, Specificity, and Accuracy of Plasma Biomarkers in Discriminating Total Hippocampal Atrophy.

| **Biomarker** | **Cutoff** | **Sensitivity / Specificity** | **Crude AUC (95% CI)\*** | **AUC (95% CI)\*\*** |
| --- | --- | --- | --- | --- |
| Covariates Only |  |  |  | 0.85 (0.71, 0.90) |
| **Singular Biomarker Model** |  |  |  |  |
| Aβ<sub>42/40</sub> | 0.38 | 0.80 / 0.57 | 0.69 (0.54, 0.84) | 0.86 (0.76, 0.96) |
| p-Tau 181 | 0.36 | 0.67 / 0.75 | 0.68 (0.51, 0.84) | 0.87 (0.78, 0.97) |
| NfL | 0.35 | 0.64 / 0.65 | 0.63 (0.48, 0.77) | 0.85 (0.75, 0.95) |
| GFAP | 0.44 | 0.50 / 0.92 | 0.73 (0.59, 0.87) | 0.85 (0.75, 0.95) |
| **Multiple Biomarkers Model** |  |  |  |  |
| Aβ<sub>42/40</sub> + p-tau 181 | - | - | 0.72 (0.55, 0.87) | 0.89 (0.80, 0.98) |
| Aβ<sub>42/40</sub> + p-tau 181 + NfL | - | - | 0.71 (0.55, 0.88) | 0.90 (0.81, 0.98) |
| Aβ<sub>42/40</sub> + p-tau 181 + GFAP | - | - | 0.78 (0.65, 0.91) | 0.89 (0.80, 0.98) |
| p-tau 181 + NfL + GFAP | - | - | 0.81 (0.70, 0.93) | 0.88 (0.79, 0.97) |
| Aβ<sub>42/40</sub> + NfL + GFAP | - | - | 0.79 (0.66, 0.91) | 0.88 (0.79, 0.97) |
| Aβ<sub>42/40</sub> + p-tau 181 + NfL + GFAP | - | - | 0.79 (0.65, 0.92) | 0.90 (0.81, 0.98) |

\* Model including only the corresponding biomarkers
\*\*Model including corresponding biomarkers and the covariates: age, gender, education, depression score, and intracranial volume

## Discussion

In the current study, we examined the discriminative ability of core AD plasma biomarkers (Aβ42/40, p-tau181) and non-specific AD biomarkers (NfL and GFAP) in predicting hippocampal atrophy in adult Congolese with and without probable AD from Kinshasa, DRC. This is one of the first exploratory studies to use AD plasma biomarkers based on Alzheimer’s Association criteria^29^ in this novel sub-Saharan African (SSA) population. Our results found that patients with AD have significantly smaller hippocampi than healthy control (HC) subjects, including both unilateral and total volumes, which is consistent with broader research literature.^30^

In our previous study, we did not find significant relationships between mood severity and hippocampal volume. Despite the lack of significant association with hippocampal volume, we used mood severity as a covariate to be conservative in our analyses. We found elevated levels of non-specific AD biomarkers (NfL and GFAP) in our patients with probable AD, which is consistent with reports that NfL and GFAP are involved in other neurodegenerative pathology.^31^

Our first hypothesis, which predicted that core (Aβ42/40 and p-tau181/217) and non-specific (NfL and GFAP) plasma biomarkers of AD would be associated with hippocampal atrophy, was partially supported. We found significant associations between p-tau 181 plasma biomarkers and lower LH volumes. However, core biomarker (Aβ42/40) and non-specific (NfL and GFAP) AD plasma biomarkers were not associated with hippocampal atrophy in this sample. These findings are similar to studies in other populations, which have found associations between AD core plasma biomarkers (p-tau 181) and hippocampal atrophy in elderly subjects with dementia.^32,33^ Only plasma p-tau181 concentrations were significantly associated with increased odds of hippocampal atrophy in this sample.

These findings also highlight the importance of core AD plasma biomarkers in the evaluation of adults with and without AD, making p-tau181 better plasma AD biomarkers in the diagnostic assessment of cognitive aging. Our results also support the utility of using these two plasma biomarkers (Aβ42/40 and p-tau181) to discriminate hippocampal atrophy in adults with and without AD in SSA populations. These core AD plasma biomarkers had greater discriminative ability for hippocampal atrophy than non-specific AD biomarkers (NfL and GFAP). These results demonstrated good specificity of plasma biomarkers in discriminating hippocampal atrophy in both crude and adjusted models. However, the sensitivity of plasma biomarkers was still poor in discriminating hippocampal atrophy.

While lateralized hippocampal findings are frequently observed in the research literature, our study did find some evidence of laterality regarding the ability of plasma biomarkers to discriminate hippocampal atrophy. The ability to discriminate atrophy was slightly better for LH than RH. The discriminative strength remained almost the same when intracranial volume was added as a covariate and after adjustment for other covariates. These results can be explained by evidence suggesting that networks in the left (i.e., dominant) hemisphere may be more vulnerable to neurodegeneration.^34^ In addition, the screening measures used for participant selection had many items that are more verbally loaded. Other studies have reported similar findings in assessing the sensitivity and specificity of plasma biomarkers in discriminating hippocampal volumes in Western elderly subjects. The findings of this study provide evidence of the usefulness of Alzheimer’s Association criteria for AD in this sample to discriminate hippocampal atrophy. Our analyses also showed the importance of MRI as a diagnostic tool for AD, with specific emphasis on hippocampal atrophy. Our findings have also demonstrated the synergistic effect of plasma biomarkers in discriminating hippocampal atrophy.

This study is the first in SSA to attempt to discriminate hippocampal atrophy based on AD core and non-specific AD plasma protein biomarkers in a novel sample of adults in the DRC with and without dementia. Our findings should be interpreted considering several limitations, such as the cross-sectional nature of the study, low sensitivity, and lack of amyloid PET imaging confirming AD pathology. These analyses should be further validated in longitudinal data. Future studies should follow a cohort to make longitudinal predictions of the rate of hippocampal atrophy based on baseline plasma biomarkers. Another limitation includes a small sample of participants, which limited the detection of differences that could have been clinically and significantly relevant to find adequate discriminative strength of the plasma biomarkers. Thus, future studies should replicate these findings with larger sample sizes. Fourth, the screening measures used (CSID and AQ) have not been validated in SSA in general and the DRC in particular. Fifth, this study may have included cases other than amnestic multidomain dementia, increasing the chances of enrolling patients with dementia caused by conditions other than AD, and thus not showing significant hippocampal atrophy. Future studies should conduct statistical analyses across all four groups (healthy controls, MCI, subjective memory complaints, and dementia). Sixth, only one brain region was analyzed. Associations with other key regions might have been identified using more powerful imaging techniques such as Voxel-Based Morphometry (VBM). Furthermore, future studies should also aim to replicate our findings using amyloid and tau brain PET or mass spectrometry to measure biomarkers. A major caveat is that our AD biomarkers were determined by Simoa, which is not optimal. The gold standard for core AD biomarker assessment is amyloid and tau brain PET. Head-to-head comparisons of amyloid and tau biomarkers have demonstrated the superiority of IPMS techniques over Simoa. Thus, continued investigation into racial disparities in AD biomarkers and their relation to AD-dementia using these gold standard techniques (e.g., brain amyloid PET, CSF) is necessary.

### Conclusions

Understanding the ability of AD core plasma biomarkers (Aβ-42/40 and p-tau 181) and non-specific plasma biomarkers (NfL and GFAP) to discriminate hippocampal atrophy in adults is a promising next step in clinical and research settings. While blood biomarkers are not equivalent to an AD diagnosis, they can be utilized as a screening tool before resorting to PET-scan neuroimaging or CSF biomarker analysis. Future longitudinal studies should test AD-related blood and CSF biomarkers from the same individuals for better discrimination of hippocampal atrophy. Additionally, larger studies with greater sample sizes and diversity in races and ethnicities should be conducted to increase generalizability.

## Data Availability

All data produced in the present study are available upon reasonable request to the authors

## AUTHOR CONTRIBUTIONS

JI: Visualization, Validation, Supervision, Software, Resources, Project administration, Methodology, Investigation, Funding acquisition, Formal analysis, Data curation, Conceptualization, Writing – review & editing, Writing – original draft. KJ: Writing – review & editing. PM: Writing – review & editing. SSP: Writing – review & editing, Writing – original draft. MS: Writing – review & editing, Writing – original draft. EE: Writing – review & editing. GG: Writing – review & editing. NT: Writing – review & editing. IK: Writing – review & editing. SM: Writing – review & editing. LM: Writing – review & editing. CT: Writing – review & editing. AS: Writing – review & editing. JR: Writing – review & editing. BC: Writing – review & editing. AL: Writing – review & editing. JK: Writing – review & editing. AB: Writing – review & editing. AJ: Writing – review & editing, Data curation. AG: Writing – review & editing. AA: Writing – review & editing.

## ACKNOWLEDGEMENTS

JI was supported by Alzheimer’s Association Research Grant (AARG)

MS was supported by the National Center for Advancing Translational Sciences of the National Institutes of Health under Award Number UL1TR002378. The content is solely the responsibility of the authors and does not necessarily represent the official views of the National Institutes of Health.

JRC was supported by AlzBio and the John Douglas French Alzheimer’s Foundation. We would like to acknowledge Dr. Allan Levey for his reading and suggestions.

